# Immunologic response, Efficacy, and Safety of Vaccines Against COVID-19 Infection in Children and Adolescents Aged 2 – 21 years old: A Systematic Review

**DOI:** 10.1101/2022.01.11.22269113

**Authors:** Sara Sadeghi, Yasamin Kalantari, Sima Shokri, Morteza Fallahpour, Nahid Nafisi, Azadeh Goodarzi

**Affiliations:** Department of Pediatrics, Division of Dermatology, University of Calgary, Alberta Children’s Hospital, Calgary, AB, Canada; Department of Dermatology, Razi Hospital, Tehran University of Medical Sciences, Tehran, Iran; Department of Pediatrics, Allergy and Clinical Immunology, Rasool Akram Medical Complex Clinical Research Development Center (RCRDC), School of Medicine, Iran University of Medical Sciences, Tehran, Iran; Department of General Surgery, Rasool Akram Medical Complex Clinical Research Development Center (RCRDC), School of Medicine, Iran University of Medical Sciences, Tehran, Iran; Department of Dermatology, Rasool Akram Medical Complex Clinical Research Development Center (RCRDC), School of Medicine, Iran University of Medical Sciences, Tehran, Iran; Skin and Stem Cell Research Center, Tehran University of Medical Sciences, Iran Jordan Dermatology and Hair Transplantation Center, Tehran, Iran

**Keywords:** COVID-19, Vaccination, Immunization, Children, Adolescents, Systematic review

## Abstract

**Backgrounds:** Children and adolescents form a large proportion of societies and play an important role in the transmission of COVID-19. On the other hand, their education, mental and physical wellness, and safety are compromised which makes vaccination a crucial step to return to normal life.

**Aims and objectives:** To collect and summarize all data about the immune response, effectiveness, and safety of available COVID-19 vaccines for people aged 2 – 21 years old.

**Methods:** A thorough systematic review was performed on available publications in English language regarding immunogenicity, efficacy, and safety of COVID-19 vaccines in individuals aged 2 – 21 years old. Types of selected studies were clinical trials, observational studies, and cohort. Searched databases comprised Ovid Medline, Cochrane Library, Embase, Scopus, Web of Sciences, Google Scholar, and ClinicalTrials.gov website. Data extraction and analysis were performed in Microsoft Word software, version 16.56.

**Results:** The COVID-19 vaccination was evaluated in a total of 50,148 children and adolescents in 22 published studies and 5,279 participants in two ongoing clinical trials. Data were collected about multiple vaccines including BNT162b2 (Pfizer), mRNA-1273 (Moderna), JNJ-78436735 (Johnson and Johnson), CoronaVac (Sinovac), BBIBP-CorV (Sinopharm), adenovirus type-5-vectored vaccine, ZyCov-D, and BBV152 (COVAXIN). The immune response and efficacy of such vaccines were 96% – 100% in healthy children and adolescents and were also acceptable in those with underlying diseases and suppressed immune systems. The current systematic review revealed favorable safety profiles of employed vaccines in children and adolescents; however, adverse reactions such as myocarditis and myopericarditis were reported which were transient and resolved entirely.

**Conclusion:** Vaccinating children and adolescents aged 2 – 21 years old is beneficial to abort the COVID-19 pandemic. Moreover, the risk-benefit assessments revealed favorable results for vaccinating children and adolescents, especially those with underlying disease, alongside adults to prevent transmission, severe infection, negative outcomes, and new variants formation.

## 1. Introduction

Severe acute respiratory syndrome coronavirus 2 (SARS-CoV-2) is caused by coronavirus 2019 (COVID-19) and was announced as a global pandemic on March 11, 2020^(1)^. Children and adolescents are at risk of COVID-19 infection as likely as other age groups; however, children may manifest milder symptoms than adults^(2)^. Although the clinical course of COVID-19 infection is smoother in pediatrics, the disease can escalate to severe pulmonary involvement especially in those with underlying medical conditions^(3)^.

Multiple trials evaluated the efficacy and safety of vaccines against COVID-19 in healthy grown-ups as well as adults with comorbidities^(4-9)^. Likewise, vaccination against coronavirus can prevent serious outcomes or hospitalization following the natural infection^(10)^. Of note, children and adolescents have their education, safety, mental and physical wellness negatively affected which it makes vaccination crucial for them^(11)^.

All children and adolescents should be considered for COVID-19 vaccination for their own protection against the infection and its outcomes, and more importantly because they are part of the COVID transmission cycle^(12, 13)^. Several clinical trials supported the favorable immune response, effectiveness, and safety profiles of COVID-19 vaccines in healthy children and adolescents, and those with underlying medical conditions^(14-16)^. In the current study, we aimed to collect all the data about immunogenicity, efficacy, and safety of available COVID-19 vaccines to guide health care workers and families on vaccinating the younger population (2 – 21 years old).

## 2. Method and Materials

The current systematic review was conducted according to the Preferred Reporting Items for Systematic reviews and Meta-Analysis (PRISMA).

### 2.1. Search Strategy

A systematic search was performed on databases including Ovid Medline, Cochrane Library, Scopus, Web of Sciences, Embase, Google Scholar, and ClinicalTrials.gov website until December 7^th^, 2021. The combination of employed keywords and MeSH terms is attached in the supplementary data (Table S1). A total of 9,369 publications were found in the primary search and 5,540 duplicates were removed in the first screening phase and 3,829 published studies entered the next phase of screening. PRISMA flow diagram can be found in supplementary data (Figure S1).

### 2.2. Literature Screening

For the entire screening process, EndNote software, version 20, was utilized. In the first phase of screening, 5,540 duplicates were detected by EndNote and removed. In the second phase of screening, two investigators independently reviewed all the literature by reading titles and abstracts to ensure their quality to be included in data extraction, and remained duplicates were removed manually. Disagreements were resolved with discussion or the consensus of the corresponding investigator. In the last phase of screening, full texts were reviewed by one investigator and 22 publications plus two ongoing clinical trials, with released interim results, were selected for data extraction.

### 2.3. Inclusion and Exclusion Criteria

Criteria for inclusion of studies comprised full text, English language, human studies, pediatric and adolescent population (21 years old or younger), clinical trials, observational studies, cohort, case series and case reports. Further, criteria excluding studies out of the review included trials about adults (older than 21 years old), studies on animals or *in vitro/ex vivo*, reviews, consensus, or guidelines, and articles which were not about COVID-19 vaccination.

### 2.4. Data Extraction

Extracted data from studies that were included in the current review are (i) study characteristics (author, year, design of study, county, name, and type of the vaccine), (ii) participants characteristics (age, sample size, and underlying medical conditions), and (iii) results (immune response, efficacy, safety, and adverse reactions). Microsoft Word software, version 16.56, was utilized for data extraction. One investigator performed data collection process.

### 2.5. Bias Assessment and Quality Evaluation

Methodological quality of the included studies and risk of bias were independently assessed by two investigators. For these assessments, the National Institute of Health (NIH) Quality Assessment Tool for Observation Cohort and Cross-Sectional Studies^(17)^ and the NIH Quality Assessment Tool for Clinical Trials^(18)^ were utilized and the results can be found in supplementary data table S2.

### 2.6. Data Analysis

We systematically reviewed and described all the information about immunogenicity, efficacy, and safety of all available vaccines for healthy children and adolescents or those with underlying diseases in Microsoft Word software, version 16.56.

## 3. Results

A total of 3,829 publications remained after removing duplicates. Among these numbers, 3,616 studies did not meet the inclusion criteria and were excluded out from the study. A total of 207 publications were entered the last phase of screening and ultimately 22 studies were selected for data extraction. Meanwhile, two ongoing clinical trials with the released interim results met the criteria to be included. Consequently, data extraction revealed the results of vaccination of a total of 50,848 children and adolescents for the current systematic review.

### 3.1. Immunogenicity of COVID-19 Vaccines in Pediatrics and Adolescents

A total of 12 publications plus two ongoing clinical trials investigated the immunogenicity of COVID-19 vaccines in participants aged 2 - 21 years old. Two other studies extended the age of their participants to 26 and 26.8 years old while assessing the immunologic response of the vaccination in pediatric groups ^(19, 20)^. Ali and Berman et al.^(21)^ reported 98.8% serologic response to mRNA-1273 (Moderna) vaccine in contributors aged 12 – 17 years old compared to 98.6% seroresponse in younger adults, and neutralizing antibodies (Ab) titers implied no inferiority in younger ages than in older. Frenck et al.^(22)^ conducted a randomized clinical trial (RCT), studying the effect of BNT162b2 (Pfizer) in participants aged 12 – 15 years old and found a greater post-vaccination Ab titer compared to vaccinated younger adults and control group. Han et al.^(23)^ Also announced over 96% serologic response 28 days after two doses of CoronaVac (Sinovac) injection to individuals aged 3 – 17 years old. Meanwhile, they noticed a higher Ab detection with 3.0μg dose of vaccine injection than 1.5μg dose. Walter ^(24)^ revealed that 99.2% of Pfizer recipients aged 5 – 11 years old achieved serologic response a month after the second dose injection. Moreover, a study conducted by Xia et al.^(25)^ on the effect of BBIBP-CorV (Sinopharm) among participants aged 3 – 17 years old reported 100% serologic response 56 days after vaccination. Noteworthy that produced Ab following the injection of 4μg and 8μg of Sinopharm were significantly higher than 2μg dosage. Furthermore, Zue et al.^(26)^ enrolled an RCT about recombinant adenovirus type-5(Ad5)-vectored COVID-19 vaccine which revealed 98% - 100% immunologic response 84 days post-vaccination in the 6 – 17-year-old age group. The robust Ab response to Ad5-vectored vaccine was higher in pediatrics than in adults.

Interim results of an ongoing RCT (*NCT04918797*) on BBV152 (COVAXIN) revealed over 90% serologic response following vaccination in 2 – 18 years old contributors. Another ongoing RCT (*NCT04796896*) has been evaluating Moderna vaccine in 4,753 individuals aged 6 months – 12 years old, and the interim results reported 99.3% immunologic response one month after the second shot of vaccine.

#### 3.1.2 Immunogenicity of COVID-19 Vaccines in Pediatrics and Adolescents with Underlying Conditions

Multiple studies evaluated the immunologic response to COVID-19 vaccines in pediatrics and adolescents with underlying clinical conditions as well as in healthy individuals. Amodio et al.^(27)^ in a case series of 21 patients, reported the effect of Pfizer vaccine in eight adolescents aged 16 – 21 years old, seven patients with common variable immunodeficiency (CVID), and one patient with unclassified Ab deficiency (unPAD). The serologic response in such patients after two doses of vaccine was significantly lower than in healthy individuals. Dailey et al.^(19)^ compared the serologic response of the natural COVID-19 infection to the immunogenicity of the several COVID-19 vaccines in Inflammatory bowel disease (IBD) patients. All patients in the latter study were under the treatment with infliximab, vedolizumab, or methotrexate and the employed vaccines were Pfizer, Moderna, and Johnson & Johnson (JNJ). The serologic response was 10 folds greater post-COVID vaccination compared to natural COVID-19 infection. In another study on adolescents with IBD, Spencer et al.^(28)^ observed that Moderna recipients developed a greater Ab response compared to Pfizer and JNJ recipients disregarding the type of immunosuppressant medication. Shire et al.^(29)^ also performed a study on 12 – 17-year-old patients with IBD treated with TNF-inhibitors. Patients received Pfizer vaccine and showed an acceptable Ab titer on follow-ups. Haskin et al.^(20)^ found 63% serorespose after two doses of Pfizer among kidney transplant recipients (KTRs) aged 13.5 – 26.8 years old. Noteworthy that a high proportion of patients without an acceptable Ab response had an eGFR<30 mL/min/1.73m^2^ and formerly received rituximab. Interestingly, KTRs with a history of natural COVID-19 infection developed higher immunologic responses compared to vaccinated KTRs. Macedoni et al.^(30)^ reported an acceptable serologic response after Pfizer vaccine in 16 – 22-year-old patients with type-1 diabetes mellitus. A total of 57 of Solid organ transplants aged 12 – 18 years old in a cohort study conducted by Qin et al.^(31)^, received Pfizer vaccine while were on multiple immunosuppressants and anti-metabolites. Serologic response was reported 56.8% after the first dose and 73.3% after the second dose of vaccine. Revon-Riviere et al.^(32)^ revealed 80% and 90% seropositive response in patients with solid tumor malignancy after the first and second dose of Pfizer, respectively. The age of patients ranged 16 – 21 years old and they were on chemotherapy, targeted therapy, or immunotherapy.

### 3.2. Efficacy of COVID-19 vaccines in Pediatrics and Adolescents

The efficacy of Pfizer vaccine in participants aged 5 – 11 years old was reported 90.7% after the second dose^(24)^ and in individuals aged 12 – 15 years old was 100% ^(22)^. In a study, assessing the effectiveness of Pfizer in 12 – 18—year-old adolescents, among 57 participants, only two patients were tested positive for COVID-19 infection, one patient before receiving the second dose and another one 46 days after the second dose^(31)^. In the category of Pfizer recipients with underlying medical conditions, adolescents with solid tumor malignancy did not develop COVID-19 infection after full immunization^(32)^.

Other vaccines such as Moderna, CoronaVac, and ZyCov-D were reported 93.3%, 65.5%, and 100% protection against COVID-19 infection in 12 – 19 years old participants, respectively ^(21, 33, 34)^. Additionally, studies on Sinopharm and COVAXIN (*NCT04918797*) suggested protection efficacy against COVID-19 among 2 – 18-year-old individuals^(25)^.

### 3.3. Safety of COVID-19 Vaccines in Pediatrics and Adolescents

Reported adverse reactions were mild-to-moderate and self-limiting. The most common adverse reactions following vaccination of children and adolescents comprised injection site pain and erythema, headache, fatigue, fever, and chills^(21-25, 27, 30, 32, 35)^. In the meanwhile, no allergic reactions were reported in patients with a history of allergy to PEG-asparaginase and acute lymphoblastic leukemia after receiving Pfizer vaccine^(36)^.

In 16 – 25-year-old patients who were residents of a long care facility and received Pfizer, after the first dose 83.9% and following the second dose of vaccine 74.2% of patients presented mild adverse reactions such as discomfort/agitation, nausea/emesis, diarrhea, fever, chills, headache, and injection site erythema^(37)^. Further, Pfizer was administered in patients with juvenile inflammatory arthritis (JIA) aged 16 – 21 years old and no exacerbation of JIA was reported^(38)^. Among KTRs aged 13.5 – 26.8 years old, a non-significant decrease in eGFR were reported after vaccination with Pfizer^(20)^. Increased agitation and changed seizure pattern (becoming cluster) were observed transiently in Pfizer recipients aged 12 – 15 years old with underlying neurologic conditions^(39)^. Pfizer vaccine was also injected to 12 – 17-year-old patients with mental illness and no adverse reactions were reported from patients^(40)^.

## 4. Discussion

Mass vaccination of children and adolescents against coronavirus can be the endgame for the current pandemic^(41, 42)^. Trials about the immunogenicity of mRNA vaccines (Moderna and Pfizer) against COVID-19 revealed a great humoral immunity and more interestingly cell-mediated response in adults and children^(43, 44)^. AstraZeneca, JNJ, and Novovax demonstrated a lower humoral response than mRNA vaccines^(44)^. The immune response in pediatric age groups was reported 90% - 100% which was also higher and more durable than natural COVID-19 infection^(21, 23, 24)^. Therefore, vaccination of children and adolescents is recommended.

Immunogenicity among children and adolescents with underlying conditions such as malignancy, IBD, transplant recipients, inherited immunodeficiency, and those on immunosuppressant and immunomodulator medications was revealed to be lower than healthy individuals^(20, 30, 45, 46)^. This finding can be justified because of the relative immune system suppression. However, it was still an acceptable immune response to vaccinate this group of children and adolescents as they are more prone to show more severe forms of COVID-19 disease and its negative outcomes^(46)^.

Full vaccination of people aged 16 years and older with mRNA vaccines provided over 90% and partial vaccination with such vaccines provided over 80% efficacy on protection against COVID-19 ^(8, 47-49)^. Other vaccines for adults such as virus-vectored vaccines (Ad26.COV2.S^(50)^, AZD1222^(51)^, Ad5-vectored^(52)^), inactivated vaccines (BBV152^(53)^, CoronaVac^(54)^), recombinant particles or nanoparticle^(55)^ (V-01^(56)^, Novavax^(57)^, CoVLP^(58)^) reported also a significant efficacy in protection against moderate to severe COVID-19 infection. Meanwhile, vaccination of children and adolescents was reported approximately 100% effective. Vaccination in 12 – 18-year-old participants has been decreased the rate of hospitalization due to COVID-19 and its consequences among these age groups^(10, 59)^.

The most common adverse reactions following COVID-19 vaccination in adult and pediatric age groups have been fatigue, body pain, injection site pain and erythema, headache, myalgia, nausea/emesis/diarrhea, fever, and joint pain^(35, 60-62)^. More serious adverse effects such as transient myocarditis and myopericarditis have been primarily reported in male adolescents; however, the incidence of such reactions is rare and most of the patients fully recovered without treatment^(63-65)^. Risk-benefit assessment for vaccination against COVID-19 determined an acceptable balance for vaccinating children and adolescents of both sexes^(63, 65, 66)^.

## 5. Conclusion

The current systematic review on 22 publications plus the interim results of two ongoing clinical trials about vaccinating children and adolescents aged 2 – 21 years-old that provided an overall result about the serologic response, efficacy, and safety of available vaccines. Vaccinating younger age groups can be helpful to end the current pandemic as kids have been a part of the COVID-19 transmission cycle. Moreover, broad vaccination of all age groups can help us to prevent other COVID-19 variants to be formed. The safety profiles of such vaccines are acceptable and make them great options to prevent COVID-19 infection in healthy children and adolescents or patients with underlying conditions such as malignancy.

## 6. Limitation and Recommendation

All reviewed studies about COVID-19 vaccines, especially in pediatric groups, are new and need more time to be evaluated about their long-term efficacy and safety. Further, more studies are required to assess the long-lasting immunity of such vaccines among pediatrics and the need for booster shots.

## Key Points

1. COVID-19 infection is milder in children and adolescents; however, vaccination in these age groups is needed to end the current pandemic and prevent the formation of new variants.
2. COVID-19 infection can cause catastrophic consequences in kids with underlying medical conditions.
3. COVID-19 vaccines provided great immune response and effectiveness (approximately 100%) in children and adolescents.
4. Available vaccines against COVID-19 infection for children and adolescents proved a favorable safety profile.
5. Serious adverse reactions such as myocarditis and myopericarditis are reported to be transient and most of the patients recovered without treatment or residual signs and symptoms.
6. Risk-benefit assessment of COIVD-19 vaccination in children and adolescents supported a favorable balance for vaccinating all ages and sexes.

## Data Availability

All data produced in the present study are available upon reasonable request to the authors

## Acknowledgment

The authors would like to appreciate Xurong (Rachel) Zhao, the librarian at Alberta Children’s Hospital, for her assistance in the database search. The authors would also like to express their gratitude to the authorities of Rasool Akram Medical Complex Clinical Research Development Center (RCRDC) for their technical and editorial assistance.

## Authors Contribution

Contributions to the current study are SS in the design, database search, screening publications, literature review, quality evaluation, and bias assessment, and drafting the manuscript. YK in screening publications, literature review, quality evaluation, and bias assessment, and drafting the manuscript, and AG, S. Shokri, MF, and NN in drafting, reviewing, and revising the manuscript critically for importance intellectual content. All authors have read and approved the final version to be published and agreed to be accountable for all aspects of the work. All authors agreed on the order in which their names are listed in the manuscript.

## Abbreviations

(SARS-CoV-2): Severe acute respiratory syndrome coronavirus 2
(PRISMA): Preferred Reporting Items for Systematic reviews and Meta-Analysis
(Ab): antibodies
(CVID): adenovirus type-5(Ad5), common variable immunodeficiency
(unPAD): unclassified pediatric antibody deficiency
(IBD): Inflammatory bowel disease
(JNJ): Johnson & Johnson
(KTRs): kidney transplant recipients
(JIA): juvenile inflammatory arthritis

## Ethical Approval

Not applicable.

## Transparency declaration

Authors declare that the manuscript is an honest, accurate, and transparent. No important aspect of the study is omitted.

## Patients and Public Partnership

Patients or the public were not involved in the design, or conduct, or reporting, or dissemination plans of our research.

## Data Availability Statement

All data produced in the present study are available upon reasonable request to the authors.

## Figures and Legends

**Table 1:** Characteristics of included published studies (n=22)

| Study ID | Country | Study design | Sample size | Age | Name of vaccine | Vaccine type | Immune Response | Efficacy | Adverse reactions and safety (n or %) | Special consideration |
| --- | --- | --- | --- | --- | --- | --- | --- | --- | --- | --- |
| **Alamer (35)** | Saudi Arabia | Cross-sectional | 965 | 12 – 18 y/o | BNT162b2<sup>1</sup> | mRNA | N/A | N/A | 60% reported at least 1 side effect | 10% had type 1 diabetes mellitus, sickle cell anemia or asthma |
| **Ali and Berman (21)** | USA | RCT<sup>2</sup> | 3,732 | 12 – 17 y/o | mRNA-1273<sup>3</sup> | mRNA | 98.8% serologic response | 93.3% (after second dose) | Injection site pain, headache, fatigue | None |
| **Amodio (27)** | Italy | Observational | 21 (only one adolescent entered to the current review) | 16 y/o | BNT162b2 | mRNA | Significant lower Ab<sup>4</sup> titer than healthy individual | N/A | Injection site pain | CVID<sup>5</sup> and Burkitt lymphoma in remission |
| **Bickel (37)** | USA | Observational | 31 | 16 – 25 y/o | BNT162b2 | mRNA | N/A | N/A | Mild adverse reactions (83.9% after the first dose and 74.2% after the second dose) | Long care facility residents |
| **Dailey (19)** | USA | Cohort | 33 | 2 – 26 y/o | JNJ-78436735<sup>6</sup> (n=5)<br>BNT162b2 (n=21)<br>mRNA-1273 (n=7) | Viral vector, mRNA | 15-fold higher serologic response post-vaccination compared to wild infection | N/A | N/A | IBD<sup>7</sup> receiving infliximab or vedolizumab |
| **Dimopoulou (38)** | Greece | Observational | 21 | 16 – 21 y/o | BNT162b2 | mRNA | N/A | N/A | Injection site reaction (74%), urticaria, no exacerbation of JIA<sup>8</sup> | JIA controlled with TNF inhibitor at least for one year |
| **Frenck (22)** | USA | RCT | 2,260 (1,131 received vaccine, 1,129 received placebo) | 12 – 15 y/o | BNT162b2 | mRNA | Greater response in adolescents than in younger adults | 100% after 2 doses, 3 cases of Covid between the first and second dose | Injection site pain, fatigue, headache, and fever | None |
| **Han (23)** | China | RCT | 552 | 3 – 17 y/o | CoronaVac (Sinovac) | Inactivated virus | Over 96% of serologic response after both doses | N/A | Injection site pain (13%), fever (25%) | None |
| **Haskin (20)** | Israel | Observational | 38 | 13.5 – 26.8 y/o | BNT162b2 | mRNA | 63% serologic response after both doses. A high proportion of patients with GFR<sup>9</sup> <30 or previously treated with rituximab did not develop Ab | N/A | Injection site reaction, fever, fatigue, headache, non-significant decrease in GFR after vaccination | Kidney transplant recipients |
| **Jara (33)** | Chile | Cohort | 38,225 (8,192 received 1 dose and 30,033 received both doses) | 16 – 19 y/o | CoronaVac | Inactivated virus | N/A | 65.5% prevents of covid-19 infection, 87.5% of hospitalization, 90.3% of ICU admission, and 86.3% of covid-related death | N/A | None |
| **King (39)** | UK | Observational | 27 | 12 – 15<br>y/o | BNT162b2 | mRNA | N/A | N/A | Severe fatigue and discomfort combined with increased agitation, change in seizure type becoming clusters | Neurologic conditions |
| **Macedoni (30)** | - | Observational | 20 | 16 – 22<br>y/o | BNT162b2 | mRNA | Acceptable serologic response | N/A | Injection site reaction and pain, fever | Type 1 diabetes mellitus |
| **Mark (36)** | Canada | Cohort | 32 | 12 – 17<br>y/o | BNT162b2 | mRNA | N/A | N/A | No allergic reactions | History of acute lymphoblastic leukemia and allergy to PEG<sup>10</sup>-asparaginase |
| **Moeller (40)** | USA | Observational | 33 | 12 – 17<br>y/o | BNT162b2 | mRNA | N/A | N/A | No adverse effects were reported from patients | Mental illness |
| **Qin (31)** | USA | Cohort | 57 | 12 – 18<br>y/o | BNT162b2 | mRNA | Ab titers 56.8% positive after the first dose and 73.3% positive after the second dose | 2 patients tested positive for mild Covid-19 (the first infected between 2 doses, the second 46 days after second dose) | N/A | Solid organ transplant recipients on multiple immunosuppressants and anti-metabolites |
| **Revon-Riviere (32)** | France | Cohort | 13 (3 patients did not receive the second dose) | 16 – 21 y/o | BNT162b2 | mRNA | Ab titers were positive in 8/10 after the first dose and positive in 9/10 after the second dose | No patients developed Covid after immunization | Injection site pain (6), fever and chills | Solid tumor malignancy on chemotherapy, targeted therapy, or immunotherapy |
| **Shire (29, 32)** | Canada | Cohort | 42 (26 patients received second dose) | 12 – 17 y/o | BNT162b2 | mRNA | Acceptable Ab response after vaccination | N/A | N/A | IBD treated with TNF<sup>11</sup> inhibitors |
| **Spencer (28)** | USA | Cohort | 340 | ≤21 y/o | JNJ-78436735 BNT162b2 mRNA-1273 | Viral vector, mRNA | 20 Patients checked for Ab after vaccination and those received Moderna had significantly higher titer of Ab | N/A | N/A | IBD on immunosuppressor |
| **Walter (24)** | USA | RCT | 2,268 (1,517 received vaccine and 751 received placebo) | 5 – 11 y/o | BNT162b2 | mRNA | 99.2% of participants achieved serologic response 1 month after the second dose | 90.7% effective (3 cases of Covid-19 reported 7 days or more after the second dose) | Fever (1 case was severe), injection site reaction and pain (71 – 74%), severe fatigue (0.9%), headache (0.3%), chills (0.1%) | 12% of participants had obesity and 8% had asthma |
| **Xia (25)** | China | RCT | 288 (phase 1), and 720 (phase 2) | 3 – 17 y/o | BBIBP-CorV (sinopharm) | Inactivated virus | 100% serologic response on day 56 post-vaccination | Protection efficacy against Covid-19 | Moderate fever (n=32), and cough (n=22) | None |
| **Zhu (26)** | China | RCT | 150 (100 received vaccine and 50 received placebo) | 6 – 17 y/o | Ad5-vectored COVID-19 vaccine | recombinant adenovirus type-5 (Ad5)-vectored | Higher Ab titers in pediatrics than in adults, 98% - 100% serologic response after 84 days | N/A | Fever, headache, fatigue, injection site pain (overall in 82%), 3 patients had severe fever, 1 had abdominal pain | None |
| **Zydus Cadila company(34)** | India | RCT | 1,000 | 12 – 18 years-old | ZyCov-D | Plasmid DNA | N/A | 66.6% (first dose)<br>100% (third dose) | 100% | None |

<sup>1</sup>Pfizer; <sup>2</sup>Randomised Clinical Trial; <sup>3</sup>Moderna; <sup>4</sup>Antibody; <sup>5</sup>Combined Variable Immunodeficiency; <sup>6</sup>Jahson & Johnson; <sup>7</sup>Inflammatory Bowel Disease; <sup>8</sup>Juvenile Inflammatory Arthritis; <sup>9</sup>Glomerular Infiltration Rate; <sup>10</sup>Polyethylene Glycol; <sup>11</sup>Tumor Necrosis Factor

**Table 2:** Characteristics of included ongoing clinical trials with released interim results (n=2)

| Clinical trial number | Country | Study design | Sample size | Age | Name of vaccine | Vaccine type | Immune Response | Efficacy | Adverse reactions and safety (n or %) | Special consideration |
| --- | --- | --- | --- | --- | --- | --- | --- | --- | --- | --- |
| **NCT04918797** | India | Clinical trial | 526 | 2 – 18 y/o | BBV152 (COVAXIN) | Inactivated virus | Over 90% serologic response | Suggested protection like adults | Suggested safety like adults | Interim results were released |
| **NCT04796896** | USA | Clinical trial | 4,753 | 6 months – 12 y/o | mRNA-1273<sup>2</sup> | mRNA | 99.3% serologic response one month after the second dose | N/A | Mild to moderate fatigue, headache, fever, and injection site pain | Continue enrolling children 6 months to 6 y/o |

<sup>1</sup>Moderna

## References

1. Jeng MJ. Coronavirus disease 2019 in children: Current status. J Chin Med Assoc. 2020;83(6):527–33.

2. Dhanya VJ. Understanding SARS-COV-2 in children: A review. European Journal of Molecular and Clinical Medicine. 2020;7(11):1102–7.

3. Jahangir M, Nawaz M, Nanjiani D, Siddiqui MS. Clinical manifestations and outcomes of covid-19 in the paediatric population: A systematic review. Hong Kong Medical Journal. 2021;27(1):35–45.

4. Butt AA, Omer SB, Yan P, Shaikh OS, Mayr FB. SARS-CoV-2 Vaccine Effectiveness in a High-Risk National Population in a Real-World Setting. ANNALS OF INTERNAL MEDICINE. 2021;174(10):1404–+.

5. Bianchi FP, Germinario CA, Migliore G, Vimercati L, Martinelli A, Lobifaro A, et al. BNT162b2 mRNA COVID-19 Vaccine Effectiveness in the Prevention of SARS-CoV-2 Infection: A Preliminary Report. JOURNAL OF INFECTIOUS DISEASES. 2021;224(3):431–4.

6. Benning L, Tollner M, Hidmark A, Schaier M, Nusshag C, Kalble F, et al. Heterologous ChAdOx1 nCoV-19/BNT162b2 Prime-Boost Vaccination Induces Strong Humoral Responses among Health Care Workers. VACCINES. 2021;9(8).

7. Bajema KL, Dahl RM, Prill MM, Meites E, Rodriguez-Barradas MC, Marconi VC, et al. Effectiveness of COVID-19 mRNA Vaccines Against COVID-19-Associated Hospitalization - Five Veterans Affairs Medical Centers, United States, February 1-August 6, 2021. MMWR Morbidity and mortality weekly report. 2021;70(37):1294–9.

8. Baden LR, El Sahly HM, Essink B, Kotloff K, Frey S, Novak R, et al. Efficacy and Safety of the mRNA-1273 SARS-CoV-2 Vaccine. The New England journal of medicine. 2021;384(5):403–16.

9. Goldschmidt K. COVID-19 Vaccines for Children: The Essential Role of the Pediatric Nurse. Journal of pediatric nursing. 2021;57:96–8.

10. Kuehn BM. COVID-19 Vaccine Highly Effective Against Adolescent Hospitalizations. JAMA. 2021;326(20):2002.

11. Marchetti F, Tamburlini G. Other good reasons for covid-19 vaccination in pre-adolescent and adolescent populations. The BMJ. 2021;374:n2052.

12. Rodewald LE, Shen K-L, Xu B-P, Yang Y-H, Wong GW-K, Namazova-Baranova L, et al. Global Pediatric Pulmonology Alliance (GPPA) proposal for COVID-19 vaccination in children. World Journal of Pediatrics. 2021;17(5):458–61.

13. Lavine JS, Antia R, Bjornstad O. Vaccinating children against SARS-CoV-2. The BMJ. 2021;373:n1197.

14. Calcaterra G, Mehta JL, De Gregorio C, Butera G, Neroni P, Fanos V, et al. Covid 19 vaccine for adolescents. concern about myocarditis and pericarditis. Pediatric Reports. 2021;13(3):530–3.

15. Dembinski L, Vieira Martins M, Huss G, Grossman Z, Barak S, Magendie C, et al. SARS-CoV-2 Vaccination in Children and Adolescents-A Joint Statement of the European Academy of Paediatrics and the European Confederation for Primary Care Paediatricians. Frontiers in Pediatrics. 2021;9:721257.

16. Khehra N, Atwal H, Padda I, Jaferi U, Narain S, Parmar MS. Tozinameran (BNT162b2) Vaccine: The Journey from Preclinical Research to Clinical Trials and Authorization. AAPS PharmSciTech. 2021;22(5):172.

17. Health NIo. Quality Assessment Tool for Observational Cohort and Cross-Sectional Studies. 2014.

18. Health NIo. Quality Assessment Tool for clinical Trials. 2014.

19. Dailey J, Hopkins D, Grandonico K, Hyams JS, Kozhaya L, Dogan M, et al. Antibody Responses to SARS-CoV-2 After Infection or Vaccination in Children and Young Adults With Inflammatory Bowel Disease. Inflammatory bowel diseases. 2021.

20. Haskin O, Ashkenazi-Hoffnung L, Ziv N, Borovitz Y, Dagan A, Levi S, et al. Serological Response to the BNT162b2 COVID-19 mRNA Vaccine in Adolescent and Young Adult Kidney Transplant Recipients. Transplantation. 2021;105(11):e226–e33.

21. Ali K, Berman G, Zhou H, Deng W, Faughnan V, Coronado-Voges M, et al. Evaluation of mRNA-1273 SARS-CoV-2 Vaccine in Adolescents. The New England journal of medicine. 2021.

22. Frenck RW, Klein NP, Brandon DM, Kitchin N, Lockhart S, Bailey R, et al. Safety, immunogenicity, and efficacy of the BNT162B2 covid-19 vaccine in adolescents. New England Journal of Medicine. 2021;385(3):239–50.

23. Han B, Li M, Wu Z, Zhao Y, Li Q, Song Y, et al. Safety, tolerability, and immunogenicity of an inactivated SARS-CoV-2 vaccine (CoronaVac) in healthy children and adolescents: a double-blind, randomised, controlled, phase 1/2 clinical trial. The Lancet Infectious Diseases. 2021;21(12):1645–53.

24. Walter EB, Talaat KR, Sabharwal C, Gurtman A, Lockhart S, Paulsen GC, et al. Evaluation of the BNT162b2 Covid-19 Vaccine in Children 5 to 11 Years of Age. New England Journal of Medicine. 021.

25. Xia S, Zhang Y, Wang Y, Wang H, Yang Y, Gao GF, et al. Safety and immunogenicity of an inactivated COVID-19 vaccine, BBIBP-CorV, in people younger than 18 years: a randomised, double-blind, controlled, phase 1/2 trial. The Lancet Infectious Diseases. 2021.

26. Zhu F, Jin P, Zhu T, Wang W, Ye H, Pan H, et al. Safety and immunogenicity of a recombinant adenovirus type-5-vectored COVID-19 vaccine with a homologous prime-boost regimen in healthy participants aged 6 years and above: a randomised, double-blind, placebo-controlled, phase 2b trial. Clinical infectious diseases : an official publication of the Infectious Diseases Society of America. 2021.

27. Amodio D, Pighi C, Medri C, Morrocchi E, Santilli V, Giancotta C, et al. Humoral and Cellular Response Following Vaccination With the BNT162b2 mRNA COVID-19 Vaccine in Patients Affected by Primary Immunodeficiencies. Frontiers in Immunology. 2021;12:727850.

28. Spencer E, Pittman NMD, Dubinsky, Marla, Klang E. Seroconversion following SARS-CoV-2 infection or vaccination in pediatric IBD patients. Journal of Pediatric Gastroenterology and Nutrition. 2021;73(1 SUPPL 1):S404–S6.

29. Shire ZJ, Reicherz F, Lawrence S, Sudan H, Golding L, Majdoubi A, et al. Antibody response to the BNT162b2 SARS-CoV-2 vaccine in paediatric patients with inflammatory bowel disease treated with anti-TNF therapy. Gut. 2021.

30. Macedoni M, Smylie G, Bolchini A, Hajro A, Petitti A, Garancini N, et al. Safety and immunogenicity of the BNT162B2 MRNA vaccine for COVID-19 in adolescents and young adults with type 1 diabetes. Pediatric Diabetes. 2021;22(SUPPL 30):15–6.

31. Qin CXT, Ashley M., Mogul DB, Auerbach SR, Charnaya O, Danziger-Isakov LA, Ebel NH, et al. Antibody response to 2-dose SARS-CoV-2 mRNA vaccination in pediatric solid organ transplant recipients. American Journal of Transplantation. 2021.

32. Revon-Riviere G, Min V, Rome A, Coze C, Verschuur A, Ninove L, et al. The BNT162b2 mRNA COVID-19 vaccine in adolescents and young adults with cancer: A monocentric experience. European Journal of Cancer. 2021;154:30–4.

33. Jara A, Undurraga EA, Gonzalez C, Paredes F, Fontecilla T, Jara G, et al. Effectiveness of an Inactivated SARS-CoV-2 Vaccine in Chile. The New England journal of medicine. 2021;385(10):875–84.

34. Abbasi J. India’s New COVID-19 DNA Vaccine for Adolescents and Adults Is a First. JAMA. 2021;326(14):1365.

35. Alamer E, Qasir NA, Areeshi H, Algaissi A, Alhazmi A, Alamer R, et al. Side effects of covid-19 pfizer-biontech mrna vaccine in children aged 12-18 years in saudi arabia. Vaccines. 2021;9(11):1297.

36. Mark C, Gupta S, Punnett A, Alexander S, Upton J, Atkinson A, et al. Safety of administration of BNT162b2 mRNA (Pfizer-BioNTech) COVID-19 vaccine in youths and young adults with a history of acute lymphoblastic leukemia and allergy to PEG-asparaginase. Pediatric Blood and Cancer. 2021;68(11):e29295.

37. Bickel S, Harris C, Huxol H, Morton R. COVID-19 Vaccination Outcomes at a Pediatric Long-Term Care Facility. The Pediatric infectious disease journal. 2021;40(7):e281–e3.

38. Dimopoulou D, Spyridis N, Vartzelis G, Tsolia MN, Maritsi DN. Safety and tolerability of the COVID-19 mRNA-vaccine in adolescents with juvenile idiopathic arthritis on treatment with TNF-inhibitors. Arthritis & rheumatology (Hoboken, NJ). 2021.

39. King H, Deshpande S, Woodbridge T, Hilliard T, Standing J, Lewis M, et al. Initial experience of the safety and tolerability of the BNT162b2 (Pfizer-Bio-N-Tech) vaccine in extremely vulnerable children aged 12-15 years. Archives of disease in childhood. 2021.

40. Moeller KE, Meeks M, Reynoldson J, Douglass M. Implementation and Outcomes of COVID-19 Vaccinations at a Child and Adolescent Psychiatric Hospital. Journal of the American Academy of Child and Adolescent Psychiatry. 2021;60(11):1332–4.

41. Schleiss MR, John CC, Permar SR. Children are the key to the Endgame: A case for routine pediatric COVID vaccination. Vaccine. 2021;39(38):5333–6.

42. Velavan TP, Pollard AJ, Kremsner PG. Herd immunity and vaccination of children for COVID-19. International Journal of Infectious Diseases. 2020;98:14–5.

43. Hershkovitz Y, Kaufman Z, Dichtiar R, Glatman-Freedman A, Bromberg M, Keinan-Boker L. Effectiveness of bnt162b2 vaccine in adolescents during outbreak of sars-cov-2 delta variant infection, israel, 2021. Emerging Infectious Diseases. 2021;27(11):2919–22.

44. Galindo R, Chow H, Rongkavilit C. COVID-19 in Children: Clinical Manifestations and Pharmacologic Interventions Including Vaccine Trials. Pediatric Clinics of North America. 2021;68(5):961–76.

45. Crane C, Ingulli E, Phebus E. Immunologic response of mRNA SARS-CoV-2 vaccination in adolescent kidney transplant recipients. Pediatric Nephrology. 2021.

46. Charla Y, Choudhury S, Kalra M, Chopra N. COVID-19 vaccination in pediatric cancer patients: A high priority. Pediatric Blood and Cancer. 2021;68(12):e29397.

47. Thompson MG, Burgess JL, Naleway AL, Tyner H, Yoon SK, Meece J, et al. Prevention and Attenuation of Covid-19 with the BNT162b2 and mRNA-1273 Vaccines. The New England journal of medicine. 2021;385(4):320–9.

48. Haas EJ, Angulo FJ, McLaughlin JM, Khan F, Pan K, Southern J, et al. Impact and effectiveness of mRNA BNT162b2 vaccine against SARS-CoV-2 infections and COVID-19 cases, hospitalisations, and deaths following a nationwide vaccination campaign in Israel: an observational study using national surveillance data. The Lancet. 2021;397(10287):1819–29.

49. Polack FP, Marc GP, Thomas SJ, Absalon J, Gurtman A, Swanson KA, et al. Safety and efficacy of the BNT162b2 mRNA Covid-19 vaccine. New England Journal of Medicine. 2020;383(27):2603–15.

50. Sadoff J, Gray G, Vandebosch A, Cardenas V, Shukarev G, Grinsztejn B, et al. Safety and Efficacy of Single-Dose Ad26.COV2.S Vaccine against Covid-19. The New England journal of medicine. 2021;384(23):2187–201.

51. Barrett JR, Belij-Rammerstorfer S, Dold C, Ewer KJ, Folegatti PM, Gilbride C, et al. Phase 1/2 trial of SARS-CoV-2 vaccine ChAdOx1 nCoV-19 with a booster dose induces multifunctional antibody responses. Nature medicine. 2021;27(2):279–88.

52. Zhu F-C, Li Y-H, Guan X-H, Hou L-H, Wang W-J, Li J-X, et al. Safety, tolerability, and immunogenicity of a recombinant adenovirus type-5 vectored COVID-19 vaccine: a dose-escalation, open-label, non-randomised, first-in-human trial. Lancet (London, England). 2020;395(10240):1845–54.

53. Ella R, Reddy S, Jogdand H, Sarangi V, Ganneru B, Prasad S, et al. Safety and immunogenicity of an inactivated SARS-CoV-2 vaccine, BBV152: interim results from a double-blind, randomised, multicentre, phase 2 trial, and 3-month follow-up of a double-blind, randomised phase 1 trial. LANCET INFECTIOUS DISEASES. 2021;21(7):950–61.

54. Zhang Y, Zeng G, Pan H, Li C, Hu Y, Chu K, et al. Safety, tolerability, and immunogenicity of an inactivated SARS-CoV-2 vaccine in healthy adults aged 18-59 years: a randomised, double-blind, placebo-controlled, phase 1/2 clinical trial. The Lancet Infectious diseases. 2021;21(2):181–92.

55. Walls AC, Fiala B, Schafer A, Wrenn S, Pham MN, Murphy M, et al. Elicitation of Potent Neutralizing Antibody Responses by Designed Protein Nanoparticle Vaccines for SARS-CoV-2. Cell. 2020;183(5):1367–82.e17.

56. Zhang J, Hu Z, He J, Liao Y, Li Y, Pei R, et al. Safety and immunogenicity of a recombinant interferon-armed RBD dimer vaccine (V-01) for COVID-19 in healthy adults: a randomized, double-blind, placebo-controlled, Phase I trial. Emerging microbes & infections. 2021;10(1):1589–97.

57. Keech C, Albert G, Cho I, Robertson A, Reed P, Neal S, et al. Phase 1–2 Trial of a SARS-CoV-2 Recombinant Spike Protein Nanoparticle Vaccine. New England Journal of Medicine. 2020;383(24):2320–32.

58. Ward BJ, Gobeil P, Seguin A, Atkins J, Boulay I, Charbonneau P-Y, et al. Phase 1 randomized trial of a plant-derived virus-like particle vaccine for COVID-19. Nature medicine. 2021;27(6):1071–8.

59. Olson SM, Newhams MM, Halasa NB, Price AM, Boom JA, Sahni LC, et al. Effectiveness of Pfizer-BioNTech mRNA Vaccination Against COVID-19 Hospitalization Among Persons Aged 12-18 Years - United States, June-September 2021. MMWR Morbidity and mortality weekly report. 2021;70(42):1483–8.

60. Al Khames Aga QA, Alkhaffaf WH, Hatem TH, Nassir KF, Batineh Y, Dahham AT, et al. Safety of COVID-19 vaccines. Journal of medical virology. 2021;93(12):6588–94.

61. Barda N, Dagan N, Ben-Shlomo Y, Kepten E, Waxman J, Ohana R, et al. Safety of the BNT162b2 mRNA Covid-19 Vaccine in a Nationwide Setting. The New England journal of medicine. 2021;385(12):1078–90.

62. Aiano F, Campbell C, Saliba V, Ramsay ME, Ladhani SN. COVID-19 vaccine given to children with comorbidities in England, December 2020-June 2021. Archives of disease in childhood. 2021.

63. Bozkurt B, Kamat I, Hotez PJ. Myocarditis with COVID-19 mRNA Vaccines. Circulation. 2021:471–84.

64. Das BB, Kohli U, Mercer C, Chaudhuri NR, Ramachandran P, Nguyen HH, et al. Myopericarditis after messenger RNA Coronavirus Disease 2019 Vaccination in Adolescents 12 to 18 Years of Age. Journal of Pediatrics. 2021;238:26.

65. Das BB, Moskowitz WB, Taylor MB, Palmer A. Myocarditis and Pericarditis Following mRNA COVID-19 Vaccination: What Do We Know So Far? Children (Basel, Switzerland). 2021;8(7).

66. Gurdasani D, Bhatt S, Flaxman S, Ratmann O, Costello A, Denaxas S, et al. Vaccinating adolescents against SARS-CoV-2 in England: a risk-benefit analysis. Journal of the Royal Society of Medicine. 2021.

