## Supplementary material for "Immunologic response, Efficacy, and Safety of Vaccines Against COVID-19 Infection in Children and Adolescents Aged 2 – 21 years old: A Systematic Review": Table S1

Searches were conducted in Ovid Medline, Ovid EMBASE, Elsevier Cochrane Library, Web of Science, Scopus, Google Scholar, and ClinicalTrials.gov.

**Supplementary Tables 1 (S1)**

**Ovid MEDLINE(R) ALL 1946 to December 07, 2021**

| **#** | **Searches** | **Results** |
| --- | --- | --- |
| 1 | exp Child/ or "Congenital, Hereditary, and Neonatal Diseases and Abnormalities"/ or exp infant/ or adolescent/ or exp pediatrics/ or child, abandoned/ or exp child, exceptional/ or child, orphaned/ or child, unwanted/ or minors/ or (pediatric* or paediatric* or child* or newborn* or congenital* or infan* or baby or babies or neonat* or pre-term or preterm* or premature birth* or NICU or preschool* or pre-school* or kindergarten* or kindergarden* or elementary school* or nursery school* or (day care* not adult*) or schoolchild* or toddler* or boy or boys or girl* or middle school* or pubescen* or juvenile* or teen* or youth* or high school* or adolesc* or pre-pubesc* or prepubesc*).mp. or (child* or adolesc* or pediat* or paediat*).jn. | 4852505 |
| 2 | COVID-19 Vaccines/ | 7147 |
| 3 | vaccination/ or mass vaccination/ | 93804 |
| 4 | vaccin*.kf,tw. | 359007 |
| 5 | 3 or 4 | 375577 |
| 6 | exp Coronavirus/ or Coronavirus Infections/ or COVID-19/ or (covid or coronaviru* or corona viru* or ncov* or n-cov* or novel cov* or COVID-19 or COVID19 or COVID-2019 or COVID2019 or SARS-CoV-2 or SARSCoV-2 or SARSCoV2 or SARSCoV19 or SARS-Cov-19 or SARSCov-19 or SARSCoV2019 or SARS-Cov-2019 or SARSCov-2019 or severe acute respiratory syndrome coronaviru* or severe acute respiratory syndrome cov 2 or 2019 ncov or 2019ncov).kf,tw. | 223880 |
| 7 | 5 and 6 | 24716 |
| 8 | 2 or 7 | 25457 |
| 9 | 1 and 8 | 2557 |
| 10 | limit 9 to (english language and yr="2019 -Current") | 2277 |
| 11 | remove duplicates from 10 | **2201** |

**Embase 1974 to 2021 December 07**

| **#** | **Searches** | **Results** |
| --- | --- | --- |
| 1 | juvenile/ or exp adolescent/ or exp child/ or exp postnatal development/ or (pediatric* or paediatric* or child* or newborn* or congenital* or infan* or baby or babies or neonat* or pre term or preterm* or premature birth or NICU or preschool* or pre school* or kindergarten* or elementary school* or nursery school* or schoolchild* or toddler* or boy or boys or girl* or middle school* or pubescen* or juvenile* or teen* or youth* or high school* or adolesc* or prepubesc* or pre pubesc*).mp. or (child* or adolesc* or pediat* or paediat*).jn. | 5052366 |
| 2 | SARS-CoV-2 vaccine/ | 7119 |
| 3 | vaccination/ | 160890 |
| 4 | vaccin*.kf,tw. | 407945 |
| 5 | 3 or 4 | 436367 |
| 6 | COVID-19/ or SARS-CoV-2/ or coronavirinae/ or betacoronavirus/ or Coronavirus infection/ or (covid or coronaviru* or corona viru* or ncov* or n-cov* or novel cov* or COVID-19 or COVID19 or COVID-2019 or COVID2019 or SARS-CoV-2 or SARSCoV-2 or SARSCoV2 or SARSCoV19 or SARS-Cov-19 or SARSCov-19 or SARSCoV2019 or SARS-Cov-2019 or SARSCov-2019 or severe acute respiratory syndrome coronaviru* or severe acute respiratory syndrome cov 2 or 2019 ncov or 2019ncov).kf,tw. | 221201 |
| 7 | 5 and 6 | 23548 |
| 8 | 2 or 7 | 24590 |
| 9 | 1 and 8 | 2630 |
| 10 | limit 9 to (english language and yr="2019 -Current") | 2356 |
| 11 | remove duplicates from 10 | **2338** |

**Google Scholar Database until December 7, 2021**

| **ID** | **Search** |  |
| --- | --- | --- |
| #1 | ["Child"] or [h "infant"] or [^"adolescent"] or ["pediatrics"] or ["child, exceptional"] or [mh ^"child, orphaned"] or [^"minor"] or (pediatric* or paediatric* or child* or newborn* or congenital* or infan* or preschool* or (pre NEXT school*) or kindergarten* or kindergarden* or (elementary NEXT school*) or (nursery NEXT school*) or ((day NEXT care*) not adult*) or schoolchild* or toddler* or boy or boys or girl* or (middle NEXT school*) or pubescen* or juvenile* or teen* or youth* or (high NEXT school*) or adolesc* or (pre NEXT pubesc*) or prepubesc*):ti,ab,kw or (child* or adolesc* or pediat* or paediat*):so |  |
| #2 | MeSH descriptor: [COVID-19 Vaccines] explode all trees |  |
| #3 | MeSH descriptor: [Vaccination] explode all trees |  |
| #4 | (vaccin*):ti,ab,kw |  |
| #5 | #3 or #4 |  |
| #6 | [Coronavirus] or [“Coronavirus Infections”] or [“COVID-19”] or (covid or coronaviru* or corona viru* or ncov* or n-cov* or novel cov* or COVID-19 or COVID19 or COVID-2019 or COVID2019 or SARS-CoV-2 or SARSCoV-2 or SARSCoV2 or SARSCoV19 or SARS-Cov-19 or SARSCov-19 or SARSCoV2019 or SARS-Cov-2019 or SARSCov-2019 or severe acute respiratory syndrome coronaviru* or severe acute respiratory syndrome cov 2 or 2019 ncov or 2019ncov). ti,ab,kw |  |
| #7 | #5 and #6 |  |
| #8 | #2 or #7 |  |
| #9 | #1 and #8 |  |
| #10 | Limit #9 to Reviews, publications Years: 2019, 2020, 2021 and languages: English | **2000** |

**Web of Science until December 07, 2021**

| **#** | **Searches** | **Results** |
| --- | --- | --- |
|  | pediatric* or paediatric* or child* or newborn* or congenital* or infan* or baby or babies or neonat* or pre-term or preterm* or premature birth* or NICU or preschool* or pre-school* or kindergarten* or kindergarden* or elementary school* or nursery school* or day care* or schoolchild* or toddler* or boy or boys or girl* or middle school* or pubescen* or juvenile* or teen* or youth* or high school* or adolesc* or pre-pubesc* or prepubesc*  **And**: covid or coronaviru* or corona viru* or ncov* or n-cov* or novel cov* or COVID-19 or COVID19 or COVID-2019 or COVID2019 or SARS-CoV-2 or SARSCoV-2 or SARSCoV2 or SARSCoV19 or SARS-Cov-19 or SARSCov-19 or SARSCoV2019 or SARS-Cov-2019 or SARSCov-2019 or severe acute respiratory syndrome coronaviru* or severe acute respiratory syndrome cov 2 or 2019 ncov or 2019ncov  **And:** vaccin*  **Limiters:**  **Publications Years:** 2019, 2020, 2021 **Languages:** English | **2570** |

**Scopus Database until December 7, 2021**

| **ID** | **Search** |  |
| --- | --- | --- |
| #1 | ["Child"] or ["infant"] or [^"adolescent"] or ["pediatrics"] or [^"minor"] or (pediatric* or paediatric* or child* or infan* or preschool* or kindergarten* or kindergarden* not adult* or schoolchild* or toddler* or (middle school*) or pubescen* or juvenile* or teen* or youth* or (high school*) or adolesc* or prepubesc*) |  |
| #2 | [COVID-19 Vaccines] or vaccination or immunization |  |
| #3 | [Coronavirus] or [“Coronavirus Infections”] or [“COVID-19”] or (covid or coronaviru* or corona viru* or ncov* or n-cov* or novel cov* or COVID-19 or COVID19 or COVID-2019 or COVID2019 or SARS-CoV-2 or SARSCoV-2 or SARSCoV2 or SARSCoV19 or SARS-Cov-19 or SARSCov-19 or SARSCoV2019 or SARS-Cov-2019 or SARSCov-2019 or severe acute respiratory syndrome coronaviru* or severe acute respiratory syndrome cov 2 or 2019 ncov or 2019ncov) |  |
| #4 | #2 and #3 |  |
| #5 | # 4 and #7 |  |
| #6 | Limit #5 to Reviews, Publications Years: 2019, 2020, 2021 and languages: English | **150** |

**Cochrane library until December 07, 2021**

| **Searches** | **Results** |
| --- | --- |
| pediatric* or paediatric* or child* or pre-school* or kindergarten* or kindergarden* or elementary school* or nursery school* or day care* or schoolchild* or toddler* or middle school* or pubescen* or juvenile* or teen* or youth* or high school* or adolesc* or pre-pubesc* or prepubesc*  **And**: covid or coronaviru* or corona viru* or novel cov* or COVID-19 or COVID19 or COVID-2019 or COVID2019 or SARS-CoV-2 or SARSCoV-2 or SARSCoV2 or SARSCoV19 or SARS-Cov-19 or SARSCov-19 or SARSCoV2019 or SARS-Cov-2019 or SARSCov-2019 or severe acute respiratory syndrome coronaviru* or severe acute respiratory syndrome cov 2 or 2019 ncov or 2019ncov  **And:** vaccin*  **Limiters:** Publications Years: 2019, 2020, 2021 and languages: English | **110** |

**ClinicalTrials.gov until 07, 2021**

**Condition or disease:** COVID or COVID-19
**Other terms:** vaccine or vaccines or vaccination
**Study Results:** Studies with Results
**Age Group:** Child
