## Supplementary material for "Immunologic response, Efficacy, and Safety of Vaccines Against COVID-19 Infection in Children and Adolescents Aged 2 – 21 years old: A Systematic Review": Figure S1

**Identification of studies via databases and registers**

Records removed *before screening*:

Duplicate records removed (n = 5,540)

Records identified from:

Databases (n = 9,369)

Ovid Medline (n=2201)

EMBASE (n= 2338)

Web of Science (n=2570)

Cochrane (n=101)

Google Scholar (n=2009)

Scopus (n=150)

**Identification**

Records screened

(n = 3,829)

Records excluded

(n = 3,616)

Reports sought for retrieval

(n = 213)

Reports not retrieved

(n = 6)

**Screening**

Reports excluded (n=160):

Reason 1: non-English (n = 1)

Reason 2: no full text (n = 2)

Reason 3: reviews (n = 124)

Reason 4: only adverse effects (n = 33)

Reports assessed for eligibility

(n = 207)

Studies included in review

(n = 47)

Reports of included studies

(n = 22)

**Included**

*From:*  Page MJ, McKenzie JE, Bossuyt PM, Boutron I, Hoffmann TC, Mulrow CD, et al. The PRISMA 2020 statement: an updated guideline for reporting systematic reviews. BMJ 2021;372:n71. doi: 10.1136/bmj.n71
