## supplementary data table S2 for "Immunologic response, Efficacy, and Safety of Vaccines Against COVID-19 Infection in Children and Adolescents Aged 2 – 21 years old: A Systematic Review"

* CD, cannot determine; NA, not applicable; NR, not reported

| Study ID | NIH Quality Assessment Tool for Clinical Trials | | | | | | | | | | | |  |  | Quality (Total Quality Score) |
| --- | --- | --- | --- | --- | --- | --- | --- | --- | --- | --- | --- | --- | --- | --- | --- |
|  | Q1 | Q2 | Q3 | Q4 | Q5 | Q6 | Q7 | Q8 | Q9 | Q10 | Q11 | Q12 | Q13 | Q14 |  |
| Ali and Berman (2021)^(1)^ | √ | √ | √ | √ | √ | √ | √ | √ | √ | √ | √ | NR | √ | √ | 13(Good) |
| Frenck (2021)^(2)^ | √ | √ | √ | √ | √ | √ | √ | √ | √ | √ | √ | NR | √ | √ | 13(Good) |
| Han (2021)^(3)^ | √ | √ | √ | √ | √ | √ | √ | √ | √ | √ | √ | NR | √ | √ | 13(Good) |
| Walter (2021)^(4)^ | √ | √ | √ | √ | √ | √ | √ | √ | √ | √ | √ | NR | √ | √ | 13(Good) |
| Xia (2021)^(5)^ | √ | √ | √ | √ | √ | √ | √ | √ | √ | √ | √ | NR | √ | √ | 13(Good) |
| Zhu (2021)^(6)^ | √ | √ | √ | √ | √ | √ | √ | √ | √ | √ | √ | NR | √ | √ | 13(Good) |
| Zydus Cadila Company (2021)^(7)^ | CD | CD | CD | CD | CD | CD | CD | CD | CD | CD | CD | CD | CD | CD | 0 (poor) |

Q1. Was the study described as randomized, a randomized trial, a randomized clinical trial, or an RCT?, Q2. Was the method of randomization adequate (i.e., use of randomly generated assignment)?, Q3. Was the treatment allocation concealed (so that assignments could not be predicted)?, Q4. Were study participants and providers blinded to treatment group assignment?, Q5. Were the people assessing the outcomes blinded to the participants' group assignments?, Q6. Were the groups similar at baseline on important characteristics that could affect outcomes (e.g., demographics, risk factors, co-morbid conditions)?, Q7. Was the overall drop-out rate from the study at endpoint 20% or lower of the number allocated to treatment?, Q8. Was the differential drop-out rate (between treatment groups) at endpoint 15 percentage points or lower?, Q9. Was there high adherence to the intervention protocols for each treatment group?, Q10. Were other interventions avoided or similar in the groups (e.g., similar background treatments)?, Q11. Were outcomes assessed using valid and reliable measures, implemented consistently across all study participants?, Q12. Did the authors report that the sample size was sufficiently large to be able to detect a difference in the main outcome between groups with at least 80% power?, Q13. Were outcomes reported or subgroups analyzed prespecified (i.e., identified before analyses were conducted)?, Q14. Were all randomized participants analyzed in the group to which they were originally assigned, i.e., did they use an intention-to-treat analysis

| Study ID | NIH Quality Assessment Tool for Observational Cohort and Cross-sectional Studies | | | | | | | | | | | | |  | |  | | Quality (Total Quality Score) |
| --- | --- | --- | --- | --- | --- | --- | --- | --- | --- | --- | --- | --- | --- | --- | --- | --- | --- | --- |
|  | Q1 | Q2 | Q3 | Q4 | Q5 | Q6 | Q7 | Q8 | Q9 | Q10 | Q11 | Q12 | Q13 | | Q14 | |  | |
| Alamer (2021)^(8)^ | √ | √ | √ | √ | √ | √ | √ | √ | √ | No | √ | NA | √ | | √ | | 12(Good) | |
| Amodio (2021)^(9)^ | √ | √ | √ | √ | No | √ | √ | √ | √ | √ | No | No | √ | | √ | | 9(Fair) | |
| Bickel (2021)^(10)^ | √ | √ | √ | √ | No | √ | √ | No | √ | CD | No | No | √ | | No | | 6(Poor) | |
| Dailey (2021)^(11)^ | √ | √ | CD | √ | No | √ | √ | CD | √ | √ | No | No | √ | | √ | | 7(Poor) | |
| Dimopoulou (2021)^(12)^ | √ | √ | √ | √ | No | √ | √ | √ | √ | √ | √ | No | √ | | √ | | 12(Good) | |
| Haskin (2021)^(13)^ | √ | √ | √ | √ | No | √ | √ | √ | √ | √ | √ | No | √ | | √ | | 12(Good) | |
| Jara (2021)^(14)^ | √ | √ | √ | √ | No | √ | √ | √ | √ | √ | √ | No | √ | | √ | | 12(Good) | |
| King (2021)^(15)^ | √ | √ | √ | √ | No | √ | √ | CD | √ | √ | √ | No | √ | | √ | | 9(Fair) | |
| Macedoni (2021)^(16)^ | √ | √ | √ | √ | CD | √ | √ | √ | √ | √ | √ | CD | √ | | √ | | 10(Good) | |
| Mark (2021)^(17)^ | √ | √ | √ | √ | No | √ | CD | No | √ | CD | √ | NA | No | | No | | 4(Poor) | |
| Moeller (2021)^(18)^ | √ | √ | No | √ | No | √ | √ | CD | √ | No | √ | No | √ | | √ | | 7(Poor) | |
| Qin (2021)^(19)^ | √ | √ | √ | √ | No | √ | √ | √ | √ | √ | √ | No | √ | | √ | | 10(Good) | |
| Revon-Riviere (2021)^(20)^ | √ | √ | √ | √ | No | √ | √ | √ | √ | √ | √ | No | √ | | No | | 9(Fair) | |
| Shire (2021)^(20)^ | √ | √ | √ | √ | No | √ | √ | √ | √ | √ | √ | No | √ | | No | | 9(Fair) | |
| Spencer (2021)^(21)^ | √ | √ | √ | √ | No | √ | CD | √ | √ | CD | √ | NA | √ | | No | | 7(Poor) | |

* CD, cannot determine; NA, not applicable; NR, not reported

Q1: Was the research question or objective in this paper clearly stated?, Q2: Was the study population clearly specified and defined?, Q3: Was the participation rate of eligible persons at least 50%?, Q4:  Were all the subjects selected or recruited from the same or similar populations (including the same time period)? Were inclusion and exclusion criteria for being in the study prespecified and applied uniformly to all participants?, Q5: Was a sample size justification, power description, or variance and effect estimates provided?, Q6: For the analyses in this paper, were the exposure(s) of interest measured prior to the outcome(s) being measured?, Q7: Was the timeframe sufficient so that one could reasonably expect to see an association between exposure and outcome if it existed?, Q8:  For exposures that can vary in amount or level, did the study examine different levels of the exposure as related to the outcome (e.g., categories of exposure, or exposure measured as continuous variable)?, Q9: Were the exposure measures (independent variables) clearly defined, valid, reliable, and implemented consistently across all study participants?, Q10: Was the exposure(s) assessed more than once over time?, Q11: Were the outcome measures (dependent variables) clearly defined, valid, reliable, and implemented consistently across all study participants?, Q12: Were the outcome assessors blinded to the exposure status of participants?, Q13: Was loss to follow-up after baseline 20% or less?, Q14: Were key potential confounding variables measured and adjusted statistically for their impact on the relationship between exposure(s) and outcome(s)?
